# Two Stage Designs for Phase III Clinical Trials

**DOI:** 10.1101/2020.07.29.20164525

**Authors:** Dean Follmann, Michael Proschan

## Abstract

Phase III platform trials are increasingly used to evaluate a sequence of treatments for a specific disease. Traditional approaches to structure such trials tend to focus on the sequential questions rather than the performance of the entire enterprise. We consider two-stage trials where an early evaluation is used to determine whether to continue with an individual study. To evaluate performance, we use the ratio of expected wins (RW), that is, the expected number of reported efficacious treatments using a two-stage approach compared to that using standard phase III trials. We approximate the test statistics during the course of a single trial using Brownian Motion and determine the optimal stage 1 time and type I error rate to maximize RW for fixed power. At times, a surrogate or intermediate endpoint may provide a quicker read on potential efficacy than use of the primary endpoint at stage 1. We generalize our approach to the surrogate endpoint setting and show improved performance, provided a good quality and powerful surrogate is available. We apply our methods to the design of a platform trial to evaluate treatments for COVID-19 disease.

## 1 Introduction

Platform trials are increasingly used to evaluate new interventions, especially in outbreak settings. For a new disease there is often a pool of potential interventions and great interest in quickly determining which are efficacious. In such a setting, a phase III trial with aggressive futility monitoring is appealing. One method is stochastic curtailment, whereby treatments are discarded if conditional power to show a statistically significant difference by the end is less than a threshold (Lan et al. (1984)) Another approach uses a beta spending function to spend the type II error rate as aggressively as desired (Pampallona et al. (2001)). Another option is the multi-arm, multi-stage (MAMS) design, an early example of which was the Systemic Therapy in Advancing or Metastatic Prostate cancer: Evaluation of Drug Efficacy (STAMPEDE) trial eliminating arms failing to meet a minimum level of performance compared to control at interim analyses Sydes et al. (2009).

The simplest approach uses two stages. In the first stage, treatments are discarded if they show little promising signal. A treatment that graduates past the first stage continues to a second stage. The combined data from the first and second stages allow for a definitive phase III evaluation. Since the first stage only serves to discard treatments, it can be thought of as an aggressive futility analysis, thus allowing a standard final analysis of both stages that completely ignores that a first stage occurred. Figure 1 shows a schematic of a sequence of 2-stage phase III trials that can evaluate 7 interventions with 3800 patients. For comparison, a sequence of single stage trials that always use 1000 patients would fully evaluate 3 interventions and be about 4/5 of the way through the 4th intervention with the same pool of 3800 patients. We get more answers with the 2-stage trial with 2 times as many declared wins and 2.5 times as many declared losses.

**Figure 1:**
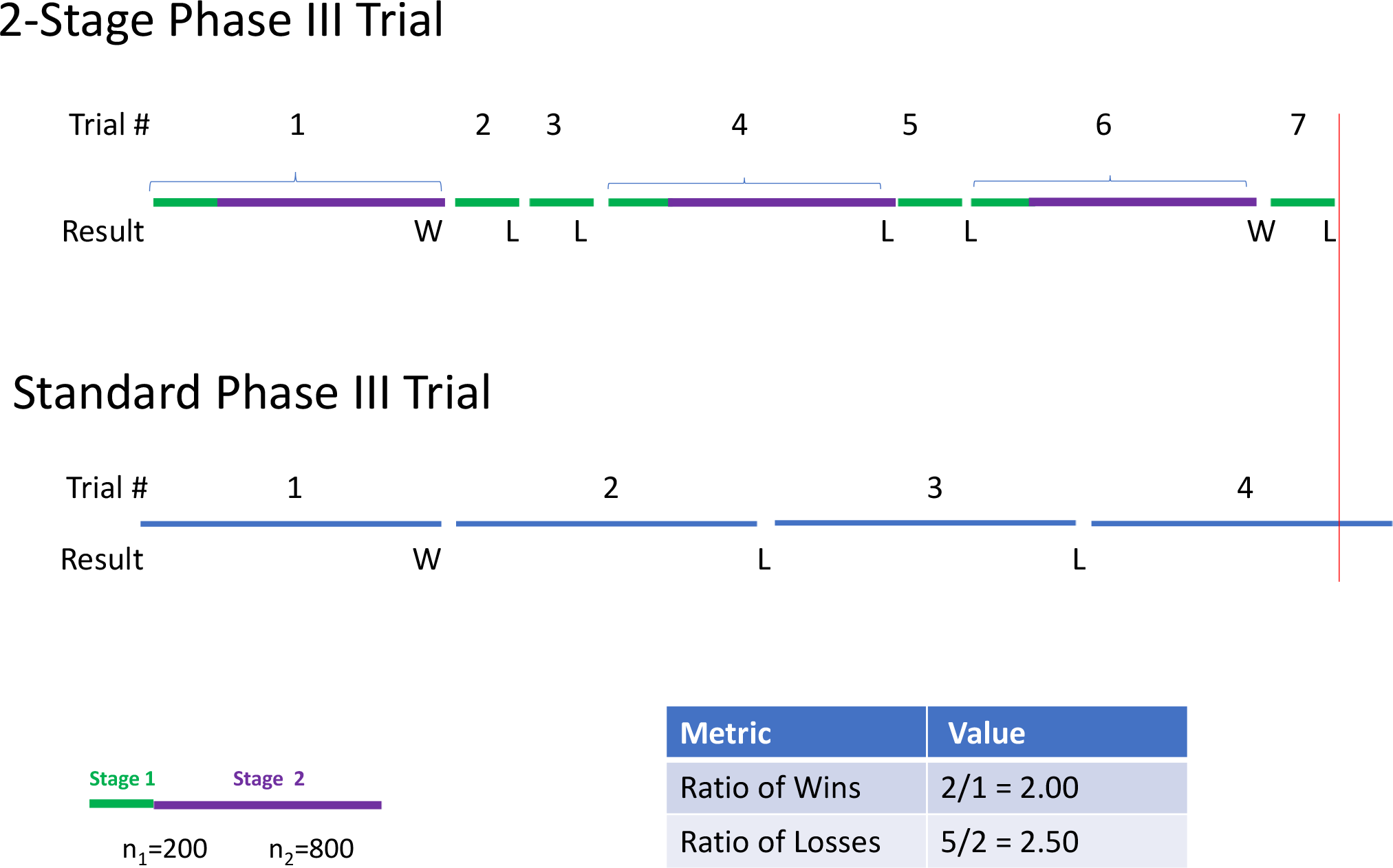
Schematic of a sequence of 2-stage phase III studies performed within a platform master protocol.

While such two stage studies have appeal, it is not clear when to define the first stage and what criteria to use to proceed to the second stage. In this paper, we evaluate how choices impact the ability to quickly determine efficacious treatments. We use Brownian Motion to approximate the test statistics over the two stages. We let *t*_1_ < 1 be the time of the stage 1 analysis and *α*_1_ be the nominal one-sided error rate for graduating from stage 1. One can imagine that each week, a fixed number of patients enroll and we evaluate the distribution of the number of identified efficacious and inefficacious treatments over the course of a year as a function of *α*_1_ and *t*_1_. We assume a fixed proportion of efficacious treatments from which we randomly select candidate treatments and design a standard trial that achieves a fixed level of power (e.g. 90%). We determine optimal values of (*t*_1_, *α*_1_) that maximize the expected number of identified efficacious treatments, relative to a standard phase III study, while maintaining high actual power(e.g. 87.5% or 89.5%).

In some settings an intermediate or surrogate endpoint might be used at stage 1. This makes sense if it is more sensitive to treatment effects than the primary and is thought to be a necessary but insufficient condition for success on the primary endpoint. Examples include early clinical readouts on a composite score portending a benefit in mortality or a good immune response to a vaccine thought necessary to cause a reduction in the incidence of disease. By specifying the power of the intermediate endpoint at trial’s end, and the correlation between the two endpoints, we can generalize our approach to this setting. We show that when using the primary endpoint, an optimal stage 1 procedure for actual power of 87.5% is around *t*_1_ = 0.40, *α*_1_ *∈* (0.28,0.49), with nearly optimal performance for (*t*_1_, *α*_1_) = (0.28, 0.49) and (0.54,0.20). This allows some flexibility in the choice of design. Use of strong surrogates at stage 1 allows an earlier stage 1 decision and substantially improves overall performance. Moderate surrogates should be avoided in favor of the primary endpoint at stage 1. We provide an example using a platform trial that evaluates monoclonal antibodies for COVID-19 disease.

## 2 Details

A large number of test statistics in clinical trials can be written as z-scores, or Wald statistics, that are approximately normally distributed when the sample size is large. At an interim analysis after fraction *t* of the trial has been completed, the z-score is denoted by *Z*(*t*). For trials with a continuous or binary outcome, the information fraction *t* is the proportion of the total planned number of patients who have been evaluated for the primary endpoint thus far. Thus, after 200 of 1000 planned patients have been evaluated, *t* = 100*/*500 = 0.20. For survival trials, *t* is the proportion of the total number of events that have occurred thus far. We can monitor clinical trials using either the z-score *Z*(*t*) or the “B-value”

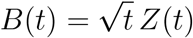

(Lan and Zucker (1993); Lan and Wittes (1988); Proschan et al. (2006)). The “B” stands for Brownian motion, a stochastic process with the following properties:

B1: For any *k* and *t*_1_ < *t*_2_ < *…* < *t*_*k*_, the increments *B*(*t*_*i*_) − *B*(*t*_*i−*1_) are independent and normally distributed, *i* = 1, *…, k. B*(0) is defined to be 0.

B2: E{*B*(*t*)} = Δ*t*.

B3: var{*B*(*t*)} = *t*.

The value Δ in property 2 is the mean of the z-score *Z*(1) at the end of the trial. If the null hypothesis is true, Δ = 0. If the alternative hypothesis is true and power is 90% for a 1-tailed test at level .025, Δ = 1.96 + 1.28 = 3.24. We can derive properties for *Z*(*t*) from those of *B*(*t*). In particular

Z1: *Z*(*t*_1_), *…, Z*(*t*_*k*_) have a multivariate normal distribution.

Z2: 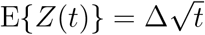

Z3: 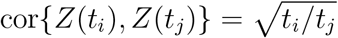 for *t_i_* ≤ *t_j_*.

At the completion of a phase III trial, we test using *Z*(1) = *B*(1), which has a standard normal distribution under the null. By properties Z1-Z3, *Z*(*t*_1_), *Z*(1) are bivariate normal with variances 1, correlation 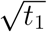, and 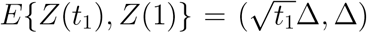. Again, Δ = 0 or 3.24 under the null or 90% powered alternative hypotheses, respectively. In this paper, we consider one-sided tests.

Our criteria for graduation from stage 1 is to reject the null that *E* {*Z*(*t*_1_)} = 0 using a test with type I error rate equal to *α*_1_. We assume that each intervention is either efficacious, *E* {*B*(1)} = Δ, which occurs with probability *θ*_1_, or inefficacious, *E* {*B*(1)} = 0, which occurs with probability *θ*_0_ = 1− *θ*_1_.

For any *a* ∈(0, 1), let *z*_*a*_ denote the (1− *a*)th quantile of a standard normal distribution. We can write the probability of declaring a winner, given that Δ is the true expected value of *Z*(1), as follows. Let *Z*_1_, *Z*_2_ be independent standard normals. Then

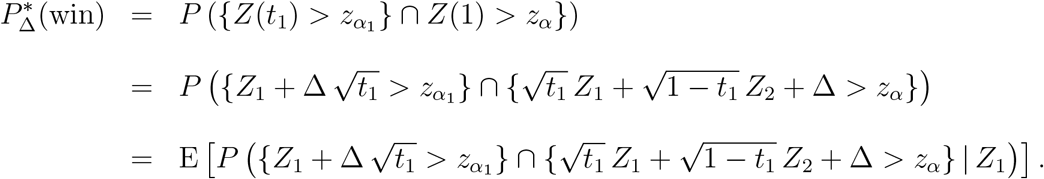

The above conditional probability given that *Z*_1_ = *z*_1_ is

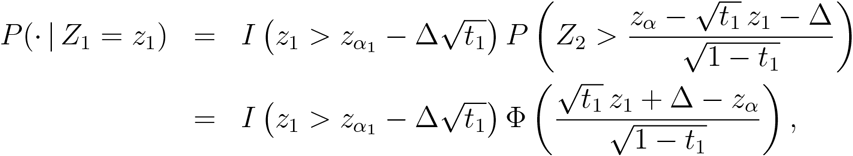

where Φ is the standard normal distribution function. Therefore, the probability of winning in a two-stage trial is

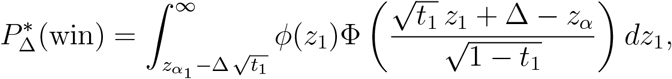

where *ϕ* is the standard normal density function. We can approximate the above integral by substituting 7 for *∞* in the upper limit of integration.

Figure 2 graphs the ‘winning’ and losing regions for *α*_1_ = 0.30. We also provide the null and alternative means for *t*_1_ = 0.30 and a bivariate normal contour to help visualize the likelihood of winning under the null and alternative.

**Figure 2:**
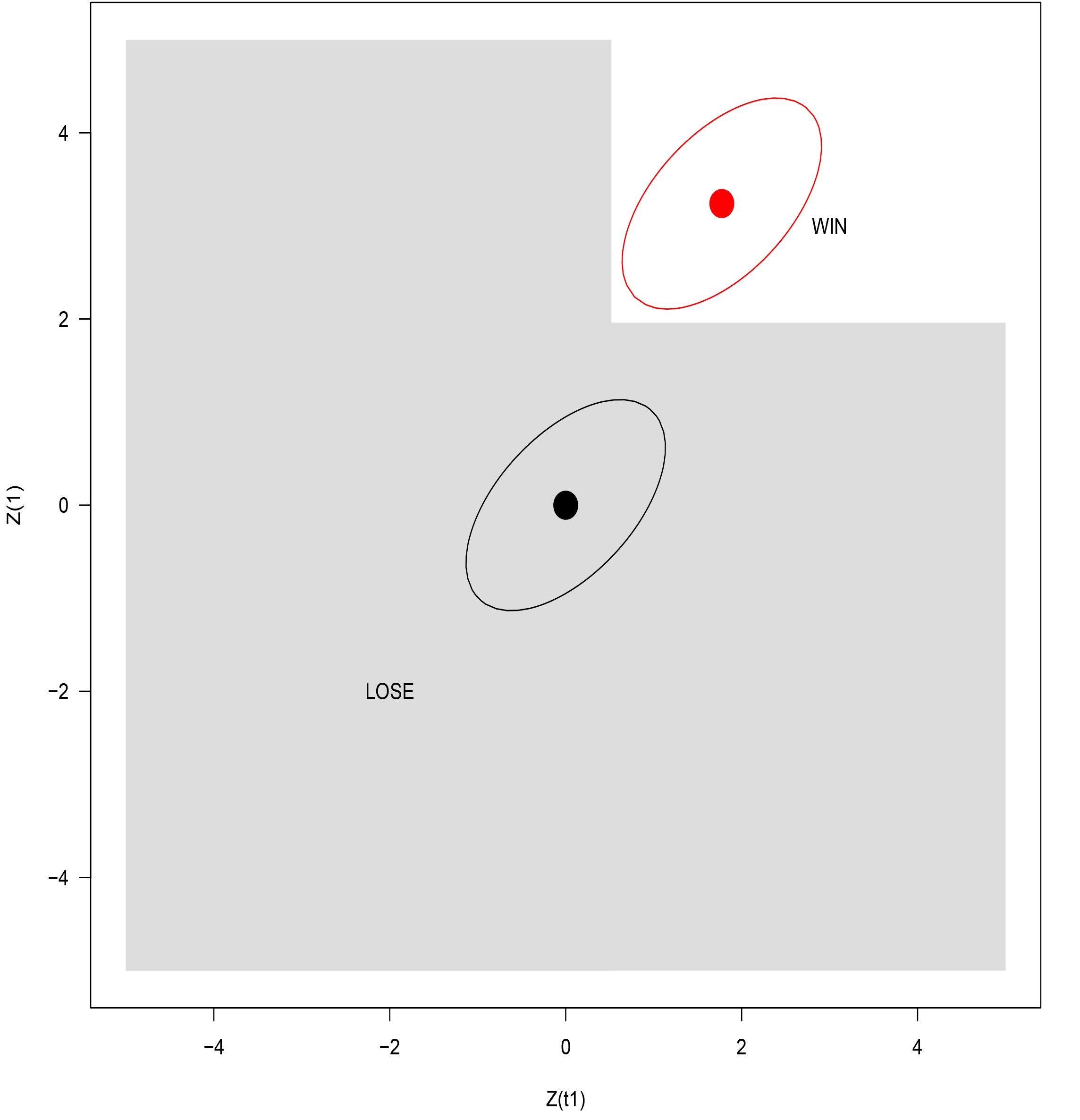
Winning and losing regions for a 2-stage trial with *α*_1_ = 0.30. For *t*_1_ = 0.30, *E* {*Z*(*t*_1_), *Z*(1) } is shown by a black circle under the null hypothesis and a red circle under the alternative hypothesis. The ellipses denote where the bivariate normal density is a constant.

The overall probability of winning is

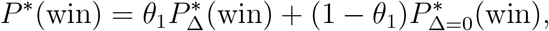

while the expected sample size is

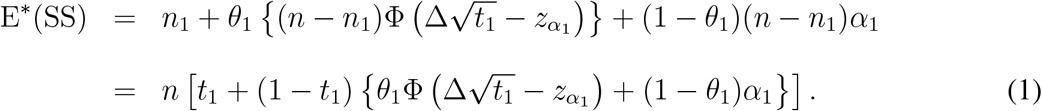

For a standard fixed sample size phase I II clinical trial, the probability of declaring a winner given Δ is

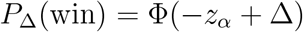

which equals 0.90 if Δ=3.24 and .025 if Δ = 0. The overall probability of winning is

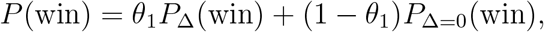

and the sample size is always *n*, thus *E*(SS) = *n*.

One way to evaluate the two-stage strategy is to evaluate the long-run expected number of interventions that are declared efficacious and inefficacious if a pool of N patients is available. To make these metrics free of time and the accrual rate, we calibrate them by dividing by the analogous quantities under the standard phase III trial. Thus, the ratio of long-run expected declared winners is

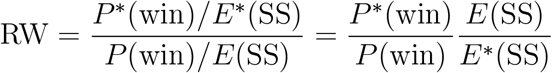

The ratio of declared losers is analogous

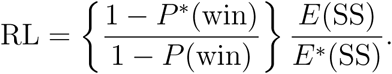

Just looking at declared winners and losers doesn’t address how often we are correct in our declarations. We know that for a standard phase III trial with *α* = .025 and *β* = .10, the probability of a false positive is 0.025 and the probability of a false negative is 0.10. With a 2-stage phase III design, the per-study false positive rate is 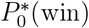 while the per study false negative rate is 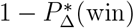. Power is 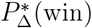. The two-stage approach will accrue fewer false positives because the criteria for success are stricter, but will have less actual power because we will incorrectly discard some true positives at stage 1.

To select good or ‘optimal’ values of (*t*_1_, *α*_1_), we would need to identify where all four metrics have good or optimal values. To simplify this problem, we focus on the ratio of wins and actual power. It would be unappealing if the 2-stage method had appreciably less power than a standard trial. Accordingly, we will fix actual power to be close to 90% and then find (*t*_1_, *α*_1_) that maximizes RW. We will also find a ‘good’ region where we achieve, say, 90% of the maximum ratio of wins.

Figure 3 graphs the RW as a function of the optimal *α*_1_ over *t*_1_ when we fix actual power at 87.5% and when half of the interventions are truly efficacious. We see that the optimal RW is 1.23, which occurs at (*t*_1_, *α*_1_) = (0.41, 0.33). The values (*t*_1_, *α*_1_) = (0.29, 0.49) and (*t*_1_, *α*_1_) = (0.54, 0.20) achieve 90% of the optimal RW. In practice, other considerations such as availability of other treatments, accrual rate, or other intermediate endpoints might come into play, thus it is reassuring that little is lost in terms of RW when allowing for flexibility in (*t*_1_, *α*_1_).

**Figure 3:**
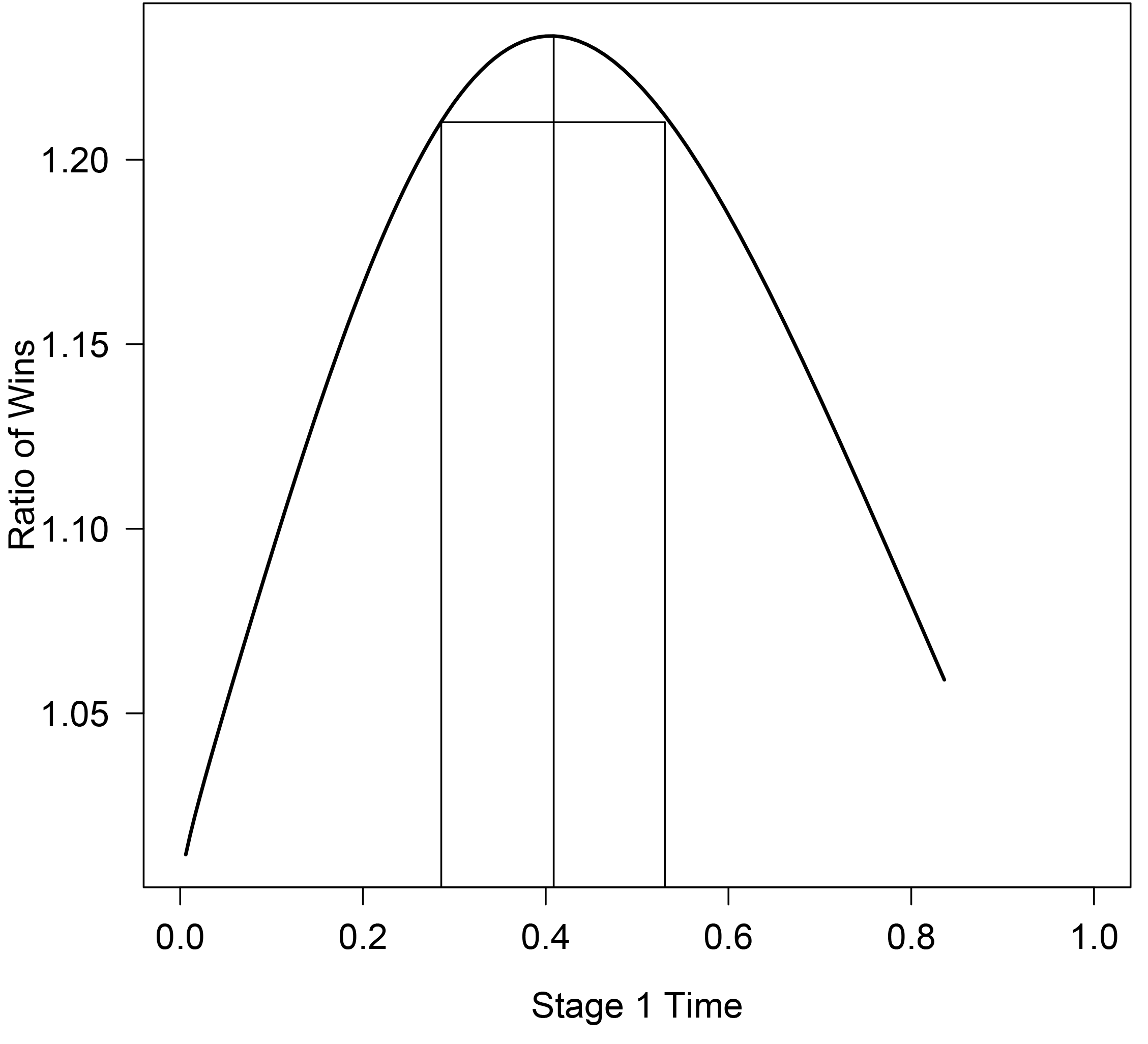
Ratio of wins (RW) as a function of *t*_1_, *α*_1_(*t*_1_) when the actual power is fixed at 87.5% and the primary endpoint is evaluated at stage 1. The maximum RW of 1.23 is achieved at *t*_1_ = 0.41, and the base of the rectangle shows values of *t*_1_ that achieve 90% of the maximum RW.

## 3 Stage 1 Surrogate Endpoint

The above development uses the same endpoint at time *t*_1_ and 1. At times, an intermediate or surrogate endpoint might make sense for the stage 1 decision. This might occur if an advantage on the surrogate endpoint was a necessary but insufficient condition for an advantage on the primary endpoint, and the surrogate endpoint was much more sensitive to treatment effects. For simplicity, we will use the term surrogate to denote any non-primary endpoint used at stage 1.

Let *Z*_*S*_(*t*) be the test statistic for a surrogate endpoint at *surrogate* information time *t*. This should accrue information at the same or faster rate for the primary endpoint. In the appendix we show that the asymptotic joint distribution of *Z*_*S*_(*t*_1_), *Z*(1) is bivariate normal with mean vector 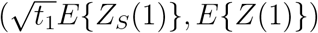, unit variances, and correlation 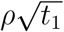. Thus, under the null, *B*_*S*_(*t*), *B*(1) is like a standard Brownian Motion but evaluated at times *ρ*^1^*t*, 1, which elegantly reflects our intuition that the correlation here should be less with use of a surrogate.

With a surrogate endpoint, the probability of winning is

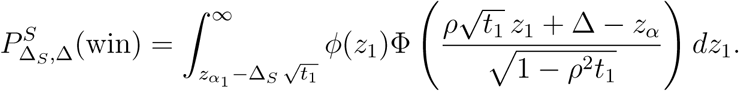

How might we specify (Δ_*S*_, Δ)? We will assume that if the surrogate shows no signal, then there is no signal on the primary, but a surrogate signal might be a false positive occasionally. In the previous section we assumed *E* {*Z*(1)} was either 0.00 or 3.24 with equal probability. Here we need to place a probability distribution on the mean vector, *E* {*Z*_*S*_(1), *Z*(1)}. We assume the possibilities are (0, 0), (0, Δ), (Δ_*S*_, 0), or (Δ_*S*_, Δ), which occur with probabilities *θ*_00_, *θ*_01_, *θ*_10_, or *θ*_11_. We assume *θ*_01_ = 0 throughout, which formalizes the idea that the surrogate is necessary but insufficient. Note that (*θ*_0_, *θ*_1_) of the previous section satisfy *θ*_0_ = *θ*_00_ + *θ*_10_ and *θ*_1_ = *θ*_01_ + *θ*_11_.

One choice for *θ*_10_ is 0 so that *E* {*Z*_*S*_(1), *Z*(1)} = (0, 0) or (Δ_*S*_, Δ). Such surrogates satisfy Prentice’s definition of surrogacy: *a response variable for which a test of the null hypothesis of no relationship to the treatment groups under comparison is also a valid test of the corresponding null hypothesis based on the true endpoint*. (Prentice (1989)).

To specify a *θ*_10_ > 0 we reason as follows. In drug development, about 60% of drugs pass phase II, and of those, about 70% pass phase III (Pretorius (2016)). Consistent with this, we can specify *θ*_00_ = 0.40, *θ*_01_ = 0.00, *θ*_10_ = (1 − 0.70) × 0.60, *θ*_11_ = 0.70 × 0.60 = (0.40, 0.00, 0.18, 0.42). To align with the previous section we will assume (0.34, 0.00, 0.16, 0.50), so that half the interventions are truly efficacious. Of course, for any specific intervention, the surrogate may be better (or worse) than this scenario, and so this choice serves as a benchmark.

With this set-up, the overall probability of a win is

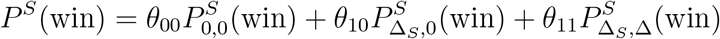

while the expected sample size is

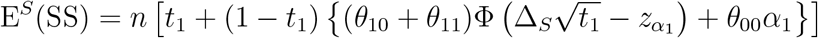

With these choices, we can evaluate a two-stage procedure that uses a surrogate endpoint, provided we estimate or specify *ρ* and *E*{*Z*_*S*_(1)}. For illustration, we specify *E*{*Z*_*S*_(1)} corresponding to either 99.5% or 95% power to formalize the idea that the surrogate should be more sensitive to treatment effects.

Table 1 summarizes the optimal and near optimal regions for various scenarios using an intermediate endpoint with final power of 99.5% or 95%, *ρ* = 0.75 or 0.10, actual power of 87.5Prentice or typical quality surrogate, respectively). The top row is the reference where we use the primary endpoint at stage 1 and set actual power at 87.5%. The optimal (*t*_1_, *α*_1_) = (0.41, 0.33). The second row changes actual power to 89.5%. This reduces RW and pushes the stage 1 look to be later with a more more liberal *α*_1_.

**Table 1:** Choices of (*t*_1_, *α*_1_) that achieve a high ratio of wins (2-stage/standard) subject to a fixed actual power. We report the optimal choice that maximizes the ratio of wins (RW) as well as choices that achieve at least 90% of the optimal RW. We evaluate settings where the primary endpoint is used at stage 1 (rows 1 and 2) as well as surrogate endpoints with varying connections to the primary endpoint. Bolded entries denote perturbations from the second row.

| $\rho$ | Surrogate Power | $\theta_{10}$ | $\theta_{11} = \text{Prop Winners}$ | Actual power | RW at Optimal | Optimal $(t_1, \alpha_1)$ | 90% of Optimal RW $(t_1, \alpha_1)$ | |
| --- | --- | --- | --- | --- | --- | --- | --- | --- |
| 1.00 | NA | NA | 0.50 | 87.5% | 1.23 | (0.41,0.33) | (0.29,0.49) | (0.53,0.21) |
| 1.00 | NA | NA | 0.50 | **89.5%** | 1.17 | (0.52,0.40) | (0.39,0.56) | (0.65,0.25) |
| 0.75 | 99.5 | 0.00 | 0.50 | 87.5% | 1.39 | (0.29,0.16) | (0.20,0.31) | (0.39,0.07) |
| 0.75 | 99.5 | **0.18** | 0.50 | 87.5% | 1.22 | (0.28,0.17) | (0.19,0.33) | (0.39,0.07) |
| **0.10** | 99.5 | 0.00 | 0.50 | 87.5% | 1.36 | (0.30,0.18) | (0.20,0.34) | (0.40,0.08) |
| 0.75 | 99.5 | 0.00 | **0.10** | 87.5% | 1.91 | (0.27,0.18) | (0.20,0.30) | (0.36,0.09) |
| 0.75 | 99.5 | 0.00 | 0.50 | **89.5%** | 1.32 | (0.37,0.21) | (0.28,0.37) | (0.48,0.10) |
| 0.75 | **95.0** | 0.00 | 0.50 | 87.5% | 1.25 | (0.38,0.32) | (0.27,0.47) | (0.50,0.20) |

The third row shows a Prentice quality surrogate which increases RW from 1.23 to 1.39, a marked improvement. The surrogate allows for an earlier look with a smaller *α*_1_ with (*t*_1_, *α*_1_) = (0.29, 0.16) instead of (0.41, 0.33). The 3rd row shows a weaker surrogate typical of those used in drug development. The optimal choice is virtually unchanged though the RW is reduced and similar to use of the primary endpoint at stage 1. The 5th row shows that the choice is robust to (within-study) correlation between the surrogate and the primary. The 6th row changes *θ*_1_ to 0.10 so that efficacious treatments are rare. The choice of (*t*_1_, *α*_1_) is virtually unchanged though RW is close to 2. This improvement in RW is because with few effiacious treatments, we discard more in stage 1, compared to *θ*_11_ = 0.50 and thus get a higher yield compared to the standard phase III design where all treatments are evaluated.

The bottom two rows show more impact on (*t*_1_, *α*_1_). The next to last row is when we require nearly 90% power which increases *t*_1_, *α*_1_. The last row shows a less powerful surrogate. The behavior here is similar to using the primary endpoint. Overall, the optimal and near optimal look times range from about 0.20 to 0.50 information with *α*s ranging from about 10% to nearly 50%.

These calculations show meaningful advantages of the 2-stage approach with little cost in terms of overall power. A variety of (*t*_1_, *α*_1_) achieve a near optimal ratio of wins and the stage 1 choice is extremely robust to changes in *θ*_10_, *ρ* and *θ*_1_. Changes in actual and surrogate power modestly change *t*_1_, *α*_1_. The calculations also highlight the importance of having a surrogate for stage 1 that does as well or better than is typical for drug development (i.e. *θ*_10_ = 0.18 or less).

## 4 Example

As a specific case, we consider the ACTIV-3 platform trial, which aims to investigate multiple therapeutics for hospitalized patients with COVID-19 disease. Approximately 1,000 patients will be enrolled in each trial to achieve 90% power using time to recovery as the primary endpoint. Interventions that show no signal shortly after randomization are thought unlikely to show benefit on recovery, and an ordinal outcome is to be measured at day 5 after randomization. Both the primary and intermediate endpoints were shown to be good choices and sensitive to treatment effects in a trial of the anti-viral remdesivir vs placebo Beigel et al. (2020).

Based on the experience in cancer trials, it was decided to use an *α*_1_ = 0.30 test at stage 1 under the expectation that the intermediate endpoint had 95% power at stage 1 with an *α*_1_ = 0.30 one-sided test with 300 patients. Thus, the trial satisfies our development with *t*_1_ = 0.30, and with 90% power for the primary endpoint, Δ = 3.24. To determine Δ_*S*_, the mean of the surrogate test statistic at trial’s end, we note that 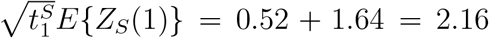, so 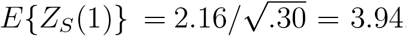 Since 1.98=3.94-1.96, we have about 98% power at the end of the study using an *α* = 0.025 one-sided test for our stage 1 endpoint. We assume our intermediate endpoint has correlation *ρ* = .75 with the primary endpoint, so 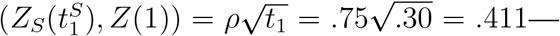 we know that performance should be robust to this choice. Finally, we assume that about half the treatments that will be evaluated are efficacious, so *θ*_11_ = 0.50 and that the intermediate endpoint is better than those typically used in drug development but worse than a Prentice surrogate. In other words *θ*_10_ should be somewhere between 0.18 and 0.00, so we specify *θ*_10_ as 0.09.

The optimal (*t*_1_, *α*_1_) is (0.35,0.28), which has a ratio of wins of 1.21 or about 20% more than standard phase III trials with fixed sample sizes. The values (*t*_1_, *α*_1_) = (0.24, 0.44) and (*t*_1_, *α*_1_) = (0.47, 0.16) preserve 90% of the optimal RW. We see that the design choices of (*t*_1_, *α*_1_) = (0.30, 0.30) are near optimal. The choice of (*t*_1_, *α*_1_) is robust to perturbations in the parameters. If we fix power at 89%, the optimum (*t*_1_, *α*_1_) goes from (0.35,0.28) to (0.42,0.32), while RW is 1.18. With a Prentice quality surrogate, i.e., *θ*_10_ = 0, the optimum (*t*_1_, *α*_1_) is unchanged but the RW goes to 1.28. With *ρ* = 0.10, the result is virtually unchanged (*t*_1_, *α*_1_) = (0.35,0.33) and RW is 1.18.

## 5 Discussion

This paper has developed a framework to evaluate performance of multiple phase III trials within a platform master protocol. We specify two stage trials, where at the end of the first stage, a decision is made whether to continue or not based on within trial outcome data. Such early stops allow for more treatments to be evaluated within a given period of time or number of patients. We evaluate performance by defining the ratio of wins (RW) for a 2-stage phase III approach and optimize the timing and ‘green light’ criteria (*t*_1_, *α*_1_) to maximize RW while fixing actual power for a given trial. We evaluate use of both the primary endpoint and a surrogate or intermediate endpoint at stage 1. With a good early stage 1 endpoint, performance can be enhanced relative to use of the primary endpoint. We find that a small decrease in actual power results in a large gain in RW under realistic parameter choices. Performance is similar for a variety of (*t*_1_, *α*_1_)s around the optimum and varies little with changes in the actual power. While we did not formally incorporate efficacy monitoring in our development, commonly used approaches such as the O’Brien-Fleming approach shoudl have little impact on performance.

There are many ways to sequentially monitor trials that allow for early stopping due to futility. The major addition of our paper is to evaluate a series of trials within a platform framework rather than a single trial. With this perspective, a new metric, RW, is a natural criterion for evaluation. In some ways, this approach is similar to Simon’s optimal design for early stage cancer studies, where a series of likely inefficacious compounds are evaluated in 2-stage one-arm studies (Simon (1989)). Potentially promising treatments are graduated for further evaluation in a second stage with expanded sample size. Thus, one can view our work as applying Simon’s perspective to phase III trials. A key difference is that under Simon’s approach, the stage 1 decision is binding, which allows a nominal *α* > 0.05 at the end of stage 2. Likewise, Magirr et al. (2012) and Ghosh et al. (2017) considered binding rules in the context of multi-arm multi-stage designs with the intent of controlling the familywise error rate across stages and arms. We view the stage 1 criteria as non-binding and feel that other information, such as results from other studies or other within-trial endpoints, can and should be allowed to over-ride the stage 1 guidance. Another contribution of our work is allowing a separate intermediate and primary endpoint. Royston et al. (2003) considered MAMS designs with a separate intermediate and definitive endpoint in stages 1 and 2, but did not unify the theory for different types of endpoints using Brownian motion with a modified information fraction. One can use Table 1 to suggest choices for a phase III trial designed with 90% power. If the primary endpoint is used at stage 1, with fixed power of 87.5% then (*t*_1_, *α*_1_) = (0.41, 0.33) is optimal, but near optimal choices range from *t*_1_=0.29 to 0.53. If the surrogate or intermediate endpoint has high power at trials end, (*t*_1_, *α*_1_) of around (0.28,0.17) is optimal and robust to surrogate connection to the primary and the expected proportion of winners in the platform trial. In practice, one could more extensively evaluate performance under a variety of scenarios to be comfortable with the design choice.

## 6 Supplementary Materials

The appendix provides a proof of that the asymptotic joint distribution of *Z*_*S*_(*t*_1_), *Z*(1) is bivariate normal with mean vector 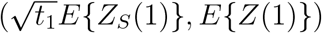, unit variances, and correlation 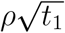.

## Data Availability

There's no data in this manuscript.

## Acknowledgments

We thank Dr. Jim Neaton, Dr. Abdul Babikar and members of the ACTIV-3 trial for motivating this work and for helpful comments.

## 7 Appendix: Surrogate and Primary Endpoint

*Theorem* Let (*X*_*i*_, *Y*_*i*_) denote the surrogate and primary endpoint for patient *i, i* = 1, *…, n*, where the *X*_*i*_ are iid with finite variance 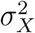 and the *Y*_*i*_ are iid with finite variance 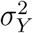. Let *ρ* = cor(*X*_*i*_, *Y*_*i*_). Consider an interim analysis at the end of stage 1 with *m* observations per arm. Let *m* →∞ and *n*→ ∞ in such a way that *m/n* → *t*. Under the null hypothesis that (***X, Y***) has the same distribution in the two arms, the joint distribution of the z-score for the surrogate endpoint at the end of stage 1 and the z-score for the primary endpoint at the end of the trial is asymptotically bivariate normal with zero means, unit variances, and correlation *ρt*^1*/*2^.

Proof. Without loss of generality, we can take E(*X*) = 0 and E(*Y*) = 0 because we can always subtract the common mean of *X*_*T*_ and *X*_*C*_, and likewise the common mean of *Y*_*T*_ and *Y*_*C*_.

Within a given arm, let

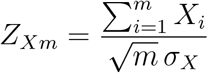

and

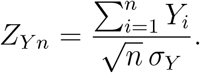

The z-scores comparing the two arms for the surrogate endpoint at stage 1 and the primary endpoint at the end of the trial are 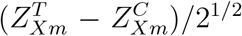 and 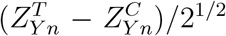, where *T* and *C* denote treatment and control.

It suffices to prove that 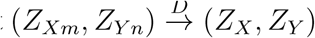, where

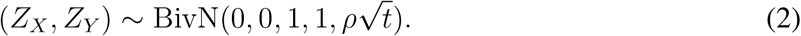

By the Cramer-Wold device, it suffices to prove that, for arbitrary constants *a* and *b, aZ*_*Xm*_+*bZ*_*Y n*_ converges in distribution to *aZ*_*X*_ + *bZ*_*Y*_. Write *aZ*_*Xm*_ + *bZ*_*Y n*_ in the following way:

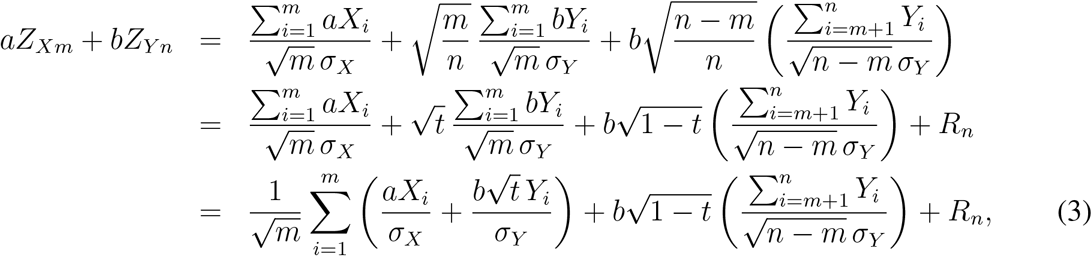

Where

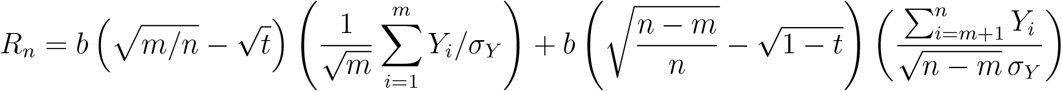

converges in probability to 0 because (*m/n*)^1*/*2^ *−t*^1*/*2^ and {(*n−m*)*/n*}^1*/*2^ *−* (1*−t*)^1*/*2^ both converge to 0 and 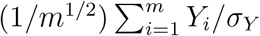 and 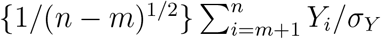 both converge in distribution to standard normals by the CLT. By Slutsky’s theorem, we can ignore *R*_*n*_ in (3). By the CLT, <u>t</u>he first term of (3) converges in distribution to a normal with mean 0 and variance 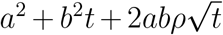, while the second term converges in distribution to a normal with mean 0 and variance *b*^2^(1− *t*). Moreover, the two terms are independent because they depend on the first *m* and last *n* − *m* iid observations, respectively. It follows that *aZ*_*Xm*_ + *bZ*_*Y n*_ converges in distribution to a normal with mean 0 and variance

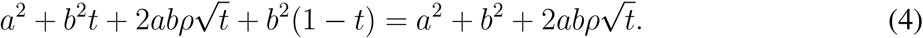

But the distribution of *aZ*_*X*_ + *bZ*_*Y*_ is also normal with mean 0 and variance given by (4). By the Cramer-Wold device, (*Z*_*Xm*_, *Z*_*Y n*_) is asymptotically normal with zero means, unit variances, and correlation *ρt*^1*/*2^, completing the proof.

